# Estimating COVID-19 Hospitalizations in the United States with surveillance data using a Bayesian Hierarchical model

**DOI:** 10.1101/2021.10.14.21264992

**Authors:** A Couture, AD Iuliano, HH Chang, NN Patel, M Gilmer, M Steele, FP Havers, M Whitaker, C Reed

**Author notes:** Corresponding Author: Alexia Couture, 1600 Clifton Road, Atlanta, GA 30329-4027. Disclaimer: The findings and conclusions in this report are those of the author(s) and do not necessarily represent the official position of the Centers for Disease Control and Prevention (CDC).

## Abstract

**Introduction:** In the United States, COVID-19 is a nationally notifiable disease, cases and hospitalizations are reported to the CDC by states. Identifying and reporting every case from every facility in the United States may not be feasible in the long term. Creating sustainable methods for estimating burden of COVID-19 from established sentinel surveillance systems is becoming more important. We aimed to provide a method leveraging surveillance data to create a long-term solution to estimate monthly rates of hospitalizations for COVID-19.

**Methods:** We estimated monthly hospitalization rates for COVID-19 from May 2020 through April 2021 for the 50 states using surveillance data from COVID-19-Associated Hospitalization Surveillance Network (COVID-NET) and a Bayesian hierarchical model for extrapolation. We created a model for six age groups (0-17, 18-49, 50-64, 65-74, 75-84, and ≥85 years), separately. We identified covariates from multiple data sources that varied by age, state, and/or month, and performed covariate selection for each age group based on two methods, Least Absolute Shrinkage and Selection Operator (LASSO) and Spike and Slab selection methods. We validated our method by checking sensitivity of model estimates to covariate selection and model extrapolation as well as comparing our results to external data.

**Results:** We estimated 3,569,500 (90% Credible Interval:3,238,000 – 3,934,700) hospitalizations for a cumulative incidence of 1,089.8 (988.6 – 1,201.3) hospitalizations per 100,000 population with COVID-19 in the United States from May 2020 through April 2021. Cumulative incidence varied from 352 – 1,821per 100,000 between states. The age group with the highest cumulative incidence was aged ≥85 years (5,583.1; 5,061.0 – 6,157.5). The monthly hospitalization rate was highest in December (183.8; 154.5 – 218.0). Our monthly estimates by state showed variations in magnitudes of peak rates, number of peaks and timing of peaks between states.

**Conclusions:** Our novel approach to estimate COVID-19 hospitalizations has potential to provide sustainable estimates for monitoring COVID-19 burden, as well as a flexible framework leveraging surveillance data.

## INTRODUCTION

Monitoring disease burden and severity is a critical component of public health research, communication, and response. The current pandemic of Coronavirus Disease 2019 (COVID-19), which is caused by Severe Acute Respiratory Syndrome Coronavirus 2 (SARS-CoV-2), has been ongoing since early 2020 and presents novel challenges and barriers to monitoring due to the unique transmission, nature of the virus, and variety of symptom presentations. In the United States, COVID-19 cases, hospitalizations, and deaths are captured through the National Notifiable Disease Surveillance System (NNDSS) and death certificates reported to the National Vital Statistics System (NVSS).^1–3^ However, hospitalization status of cases reported by states through NNDSS is often incomplete and thus might inaccurately represent the burden of COVID-19 hospitalization in the United States. In addition, since July 15, 2020, hospitalizations known or suspected to be related to COVID-19 have been reported daily through HHS Protect, known as the Unified Hospital Timeseries Data.^4^ This data collection is a burden on facilities that is likely unsustainable in the long term.

Current research and methods for estimating hospitalizations of COVID-19 are limited. In mid 2020, the Centers for Disease Control and Prevention (CDC) developed a multiplier method for estimating infections and hospitalizations of COVID-19 based on state and territory reported line-level case data.^5^ Other papers have leveraged seroprevalence surveys to estimate infections and hospitalizations of COVID-19.^6,7^ These methods rely on data systems such as case reporting or wide scale, special seroprevalence surveys that were initiated during the pandemic but might not exist in the future, as the pandemic winds down. Case-count data and consistent, representative seroprevalence data may eventually be discontinued due to the pandemic slowing down and resources and attention going elsewhere; leaving a need for longer term systems that can be sustained.

Since March 2020, the Coronavirus Disease 2019-Associated Hospitalization Surveillance Network (COVID-NET) has collected data on laboratory confirmed SARS-CoV-2-positive patients from a network of hospitals in 14 U.S. states.^8^ Although this sentinel surveillance system does not cover the entire United States, it is expected to continue monitoring rates of COVID-19 hospitalization even after the pandemic ends. The COVID-NET system was built off of the similar long-standing Influenza Hospitalization Surveillance Network (FluSurv-NET), which has been monitoring population-based rates of influenza hospitalization for almost 20 years.^9^

We created a method to utilize COVID-NET data to provide national and state-specific estimates of hospitalization to provide a long-term, sustainable framework to generate estimates of COVID-19 disease burden in the United States. The aim of this study was to estimate monthly COVID-19 hospitalization rates for all 50 states from May 2020 through April 2021. We adapted a Bayesian hierarchical model to estimate and extrapolate hospitalization rates, accounting for uncertainty and variability between states and across time.

## METHODS

### COVID-NET Surveillance Hospitalization Data and Adjustments

We used COVID-19 hospitalization data from COVID-NET. The network identifies patients who tested positive for SARS-CoV-2 through a test ordered by a health care professional and who are hospitalized within 14 days of their positive test. Hospitalization rates are calculated by the number of residents in a catchment area, defined as the area/population around the reporting hospital that the hospital potentially services, of the COVID-NET sites who are hospitalized with confirmed, positive SARS-CoV-2 test divided by the total population within that defined catchment area. The network is made up of over 250 acute-care hospitals representing 99 counties in 14 states: California, Colorado, Connecticut, Georgia, Iowa, Maryland, Michigan, Minnesota, New Mexico, New York, Ohio, Oregon, Tennessee, and Utah. Overall, the network covers about 10% of the United States population. For this analysis, case data were aggregated by month of hospitalization, state reporting, and the following 6 age groups, 0-17 years, 18-49 years, 50-64 years, 65-74 years, 75-84 years, and 85 years and older.

Recognizing that all hospital patients are unlikely to be tested for SARS-CoV-2 and, therefore, some true cases are not classified as COVID-19 patients, COVID-19 hospitalization rates are adjusted by weighting them for SARS-CoV-2 testing practices, i.e., the probability of being tested for SARS-CoV-2 during their hospitalization. In addition, testing practices changed over the course of the pandemic. The probability of being tested was calculated from IBM Watson Health Explorys electronic health record database (IBM Corporation, Armonk, NY), which includes more than 39 health system partners across the country. All states participating in COVID-NET, except Connecticut, used the same testing probabilities calculated from IBM Watson data, which were aggregated testing practices of all partners stratified by month and age group. The testing probabilities for these 13 states ranged from 0.28 to 0.67. Connecticut provided site-specific testing practice data through COVID-NET, which ranged from 0.32 to 1.00. Rates were also adjusted to account for the SARS-CoV-2 assay sensitivity because, depending on the sensitivity of the assay, some patients could have false negative test results (i.e., would not be identified as a COVID-19 hospitalization). The assay sensitivity was assumed to be 0.885, which is the midpoint for the range found in a systemic review.^10^ The adjusted hospitalization counts were used to calculate rates, using COVID-NET catchment populations for each site, i.e., as the model input rates. Due to range in hospitalizations by age groups overtime, six models were run, one for each age group.

For each age group,

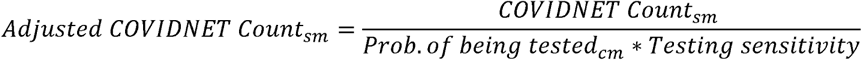

where s=1,..,S for each COVID-NET state, m=1,..,M for each month, and c= Connecticut or not Connecticut.

### Covariate Data and Selection

To extrapolate COVID-19 hospitalization rates from COVID-NET sites to states not included in the COVID-NET network, we incorporated model covariates based on state, month, and age-specific demographic and epidemiological data. We used different data measures to account for differences between states with COVID-NET sites and those states without COVID-NET sites from multiple sources (Table 1). Including covariates in the model helps to quantify differences between age groups, months, and states and allows for the model to account for these differences when estimating how many COVID-19 hospitalizations have occurred. We considered both time-varying and time-invariant state-level covariates that captured other COVID-19 disease trends, population demographics, and population health indicators. For the time varying covariates, we considered the percent of SARS-CoV-2 positive tests from commercial and public health laboratories, percent of all-cause deaths that were coded as COVID-19 deaths from National Center for Health Statistics (NCHS) and NVSS, and the following hospital capacity variables: percent COVID-19 patients out of all inpatients and percent intensive care unit (ICU) beds occupied out of all ICU beds.^11-15^ We incorporated a one week lag to the percent positive COVID-19 tests to account for time between symptom onset and hospitalization and a one week lead to the percent of COVID-19 deaths out of all deaths to account for time between hospitalization and death. For the time-invariant covariates, we considered the percent Native American and percent Black American, and the population prevalence of the following conditions/diseases from the Behavioral Risk Factor Surveillance System (BRFSS): any chronic condition, obesity, heart disease, chronic obstructive pulmonary disease (COPD), diabetes, chronic kidney disease (CKD), and asthma.^16,17^ Underlying medical and chronic conditions were found to be highly prevalent in hospitalized COVID-19 patients and were therefore included as possible covariates.^18^ Table 1 summarizes all of the variables that were considered as covariates. We used covariate selection methods to determine which of the possible covariates to include in the model. For the <18 year old age group, only asthma was included as a possible covariate from the chronic conditions/diseases because of lack of evidence that the prevalence of other chronic conditions/diseases affected COVID-19 hospitalization in that age group.

**Table 1:** Variables considered to be covariates with stratification and source

| **VARIABLES** | **STRATIFIED BY** | **SOURCE** |
| --- | --- | --- |
| **Laboratory surveillance:**<br>SARS-CoV-2 percent positive using rt-PCR tests | Month, State, Age | Commercial lab and public health lab data |
| **Vital records death:**<br>Percent of all-cause deaths that were coded as COVID-19 deaths | Month, State, Age | National Center for Health Statistics National Vital Surveillance System |
| **Hospital capacity:**<br>% COVID patients out of all inpatients, % ICU occupied out of all ICU beds | Month, State | HHS Protect /National Center for Health Statistics |
| **Race/Ethnicity:**<br>% American Indian, % Black, % Racial Minority | State, Age | National Center for Health Statistics / National Vital Statistics System |
| **Chronic conditions/diseases:**<br>% any chronic, % obesity, % heart disease, % COPD, % Diabetes, % CKD, % asthma | State | CDC MMWR Stacks / The Behavioral Risk Factor Surveillance System |

Note: Racial minority defined as non-white and non-Hispanic.

Extreme values were detected for time-varying covariates and subsequently transformed using Winsorization, i.e., minimized the influence of outliers by replacing them by the maximum and/or minimum values at a threshold of distribution percentiles.^19^ We used the adjusted COVID-NET hospitalization rates as the outcome to select covariates, separately for each age group. Covariate selection methods assist with avoiding collinearity and ensuring that the most relevant and impactful covariates are included. Our method for covariate selection utilized Least Absolute Shrinkage and Selection Operator (LASSO) and Spike and Slab.^20,21^ Covariates were included in the final model for the specific age group if they were selected by LASSO and then the model incorporated spike and slab selection. The LASSO chooses a subset of predictors by introducing an upper bound for the sum of squares and minimizing the errors present in the model. Spike and slab is a Bayesian approach where we assigned priors to the regression coefficients to be zero or non-zero, which is where the name comes from. From that, the posterior distributions show a bi-separation effect in the model coefficients – those that peak at zero and those significantly different from zero. Assumption for non-zero was high in the model due to LASSO selection being done first.

### Bayesian Hierarchical Model and Extrapolation

We implemented a Bayesian hierarchical model for extrapolation adapted from a model to estimate global influenza burden rates.^22^ Parameter estimation and inference were conducted under a fully Bayesian framework to better quantify uncertainties in predicted hospitalization rates, including those that are extrapolated to states without COVID-NET data.

We let 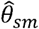 denote the estimated COVID-19 hospitalization rate, per 100,000 population from the COVID-NET states during months from the pandemic, starting in May 2020, where s=1,…,S, S=14 states in COVID-NET, m=1,…,M, and M=12 for each month included in the model. For each hospitalization estimate, the standard error and corresponding variance, 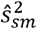, were calculated from the standard error of the rate from a Poisson distribution. Those estimated numbers, along with the selected covariates, were used as inputs into the following Bayesian hierarchical model:

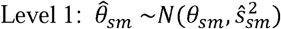

where 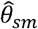 is the estimated hospitalization rate for state and month from COVID-NET data, 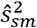 is the corresponding estimated variance, and θ_*sm*_ is the unobserved true hospitalization rate.

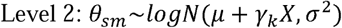

where *X* is the covariate matrix, and covariates are with mean zero and variance 1.

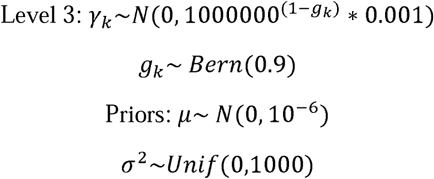

where k=1,…,K and K=the number of selected covariates.

Inference was carried out utilizing Markov chain Monte Carlo (MCMC) simulations with 20,000 iterations. The model outputs included samples from the posterior distribution of COVID-19 associated hospitalizations for each state and month. Using these samples, we calculated the median and 90% credible intervals for hospitalization counts, rounded to the hundreds due to MCMC errors, and used the state population by age group to calculate final hospitalization rates. To calculate overall age, age by month, age by state, and state by month hospitalizations and rates, we first summed the posterior samples. Since the median of sums does not equal the sum of medians, this led to slightly different total hospitalizations depending on which grouping was used to sum. For consistency, we calculated total hospitalizations from overall age medians, total monthly hospitalizations from age by month, and total state hospitalizations by age by state. We chose 20,000 iterations after starting with 2,000 iterations and slowly increasing to obtain stable estimates that also minimized simulation error.

### Validation/Comparison

We conducted sensitivity analyses to assess the effect of covariate selection and input data on the model. Multiple combinations of covariates were examined for each age group to assess how robust the hospitalization estimates were to covariate selection. To validate and test the sensitivity of the model, first, we compared how the model estimated hospitalizations for each COVID-NET state with the observed hospitalization rate from COVID-NET. In another sensitivity analysis, we dropped data from each COVID-NET state, one by one, and then compared the observed hospitalization rates to the extrapolated rates for each dropped state. Finally, we also compared our COVID-19 hospitalization estimates against other estimates and databases, including COVID-19 hospitalization rates reported through Healthdata.gov, COVID Tracking project, and from our earlier burden model, CDC’s case-based multiplier model.^5^ The Unified Hospital Timeseries data and COVID Tracking project are publicly available. According to Healthdata.gov, the Unified Hospital Timeseries data had reliable counts of new hospitalization with COVID-19 starting in the fall of 2020 when over 95% reporting from all hospitals reported by the Department of Health and Human Services (HHS). CDC’s case-based multiplier model estimates hospitalization in two month increments and by HHS regions, not state. Our model output was aggregated appropriately for comparisons.

## RESULTS

The covariates selected for each age group varied (Supplementary (S) Table 1). The SARS-CoV-2 percent positive, the percentage of COVID-19 inpatients out of all inpatients, and the percentage of hospitalizations that were ICU admissions were selected for each of the age groups. The 18-49 year old age group had the most covariates selected, and the <18 year old age group had the fewest covariates selected.

From May 2020 through April 2021 in the United States, we estimated there were 3,569,500 (90% Credible Interval (CrI): 3,238,000 – 3,934,700) hospitalizations representing a rate of 1,089.8 (90% CrI: 988.6 – 1,201.3) hospitalizations per 100,000 population. The estimated rates varied by age group, state, and month. The highest rates of hospitalization were among those aged 85 years and older, with a rate of 5,583.1 per 100,000 population (90% CrI: 5,061.0 – 6,157.5), and the lowest hospitalization rate was for those less than 18 years of age, with a rate of 80.9 per 100,000 population (90% CrI: 73.3 – 88.6). Table 2 summarizes the final estimated counts and rates of hospitalizations by age group from May 2020 through April 2021.

**Table 2:** Cumulative hospitalization count (median) and rate per 100,000 population and accompanying 90% credible intervals for each age group, and overall, from May 2020 through April 2021.

| **Age Group** | **Hospitalization Count** | **90% CrI** | **Hospitalization Rate per 100,000** | **90% CrI** |
| --- | --- | --- | --- | --- |
| <18 years | 59,000 | 53,500 – 64,600 | 80.9 | 73.3 – 88.6 |
| 18-49 years | 887,600 | 802,600 – 987,700 | 643.9 | 582.3 – 716.6 |
| 50-64 years | 924,700 | 844,100 – 1,013,200 | 1,472.0 | 1,343.7 – 1,612.9 |
| 65-74 years | 706,900 | 641,800 – 774,200 | 2,248.8 | 2,041.8 – 2,463.1 |
| 75-84 years | 623,300 | 562,500 – 689,000 | 3,909.2 | 3,527.5 – 4,320.9 |
| ≥85 years | 368,100 | 333,700 – 405,900 | 5,583.1 | 5,061.0 – 6,157.5 |
| **TOTAL** | **3,569,509** | **3,238,000 – 3,934,700** | **1,089.8** | **988.6 – 1,201.3** |

Hospitalization rates for all age groups peaked in either December 2020 or January 2021. Figure 1 shows the epidemiologic curves of hospitalizations over time by age group. During the study period, we observed the largest peak in hospitalization rates in December 2020 (183.8/100,000), followed by January 2021 (180.2/100,000). The second, smaller peak in COVID-19 hospitalizations was observed for all age groups in July 2020 (90.5/100,000). The lowest rate of hospitalization was observed across age groups in September 2020 (46.4/100,000). Following the peak in COVID-19 hospitalization rates during the winter months, COVID-19 hospitalizations declined until the month of April 2021 (Figure 1).

**Figure 1:**
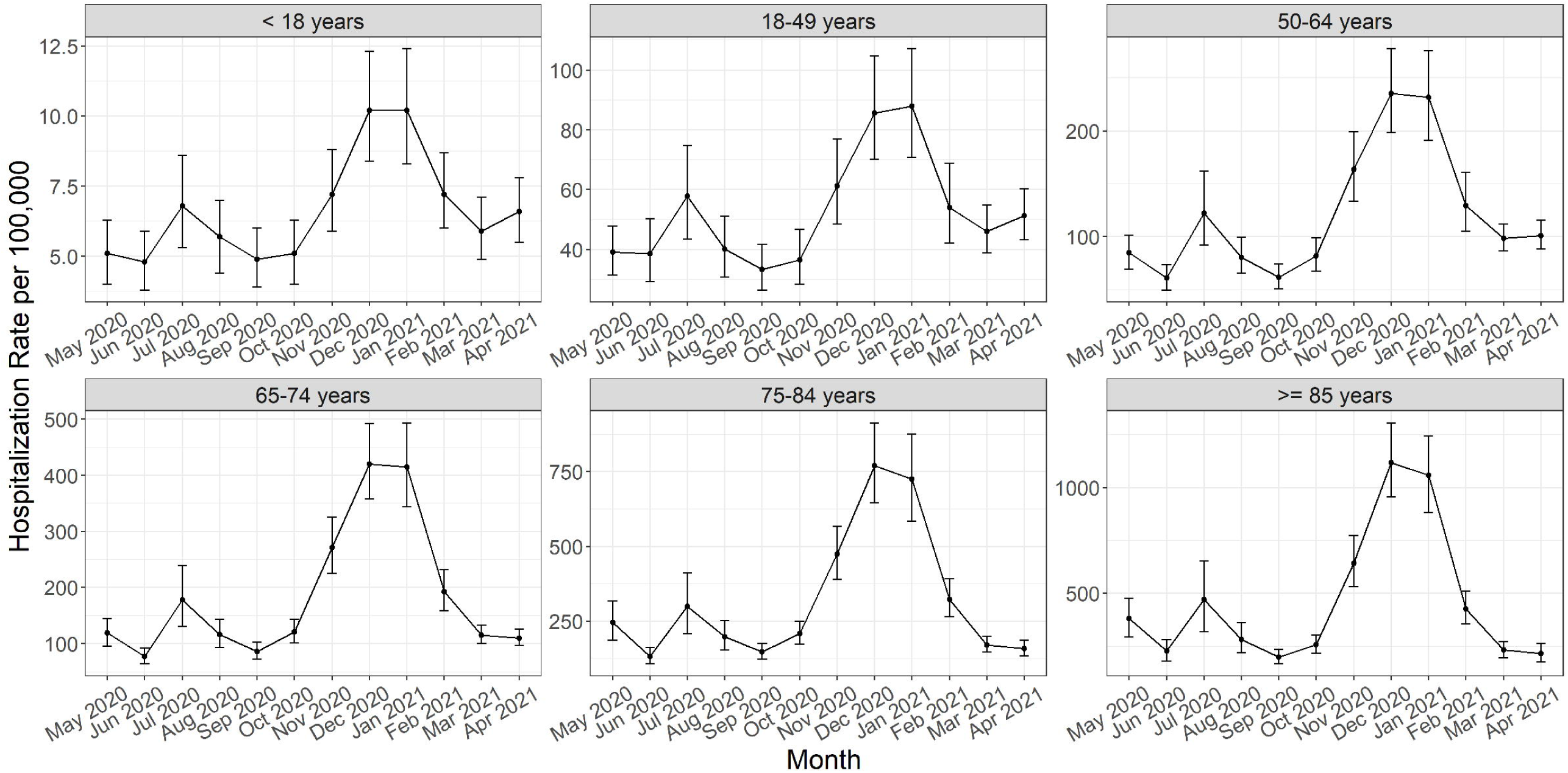
Hospitalization rates per 100,000 population and 90% credible intervals by age group over time from May 2020 through April 2021 from final model. Footnote: Y-axis limits adjust to the unique range for each age group., i.e. they are not set to the same scale.

At a state level, cumulative hospitalization rates from May 2020 through April 2020 ranged from 351.8 (90% CrI: 241.5 – 476.6) hospitalizations per 100,000 people in Vermont to 1,821.0 (90% CrI: 1,137.5 – 2,592.8) hospitalization per 100,000 people in Nebraska. Figure 2 shows the overall cumulative hospitalization rate per 100,000 people from May 2020-April 2021 for all states with a heat map (panel a) and by bar graph (panel b) to show in the range of hospitalization burden across the country. COVID-NET states are well-distributed throughout the highest to lowest rates by state (panel b).

**Figure 2:**
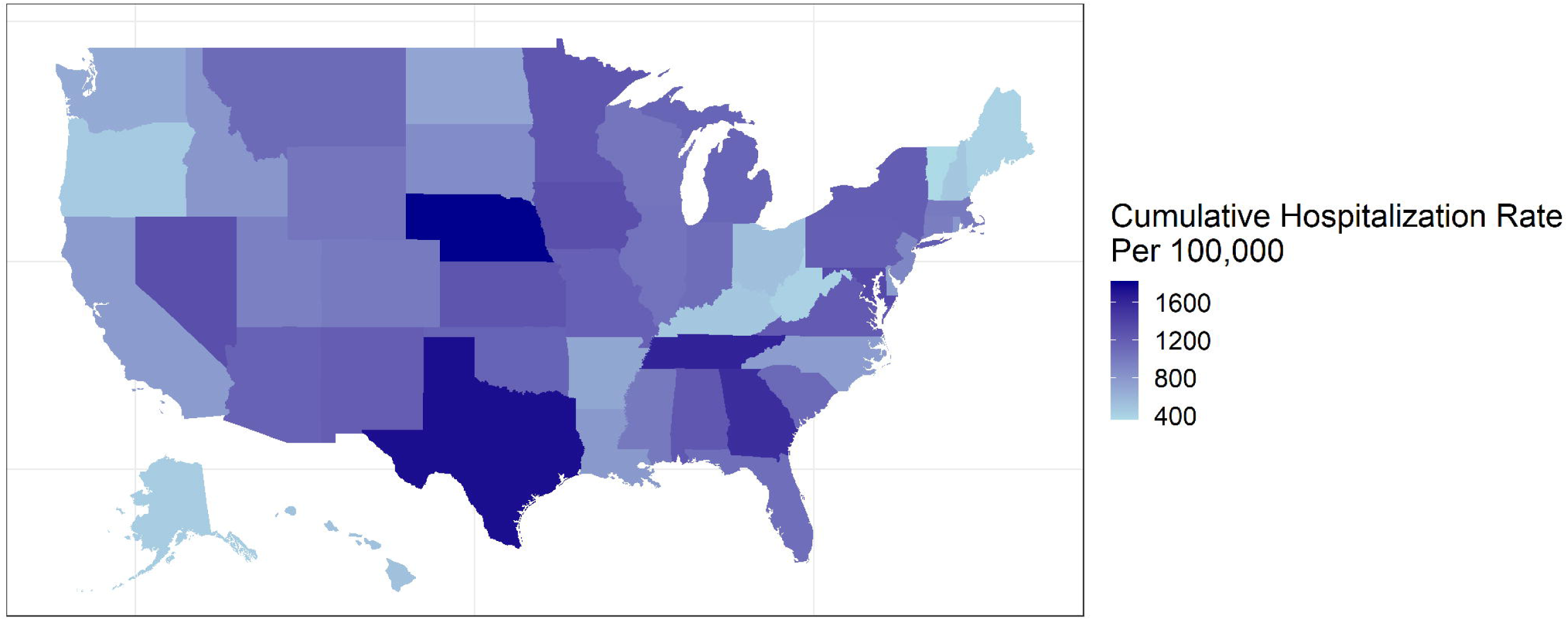
Cumulative hospitalization rate per 100,000 population by state from May 2020 through April 2021. Panel a: Heat map of the United States of cumulative hospitalization rate per 100,000 population from May 2020 through April 2021. Panel b: Bar chart of cumulative hospitalization rate per 100,000 population from May 2020 through April 2021 with 90% credible intervals with states from COVID-NET in blue. Footnote: The color range in panel a is from 352 (lightest) to 1,821 (darkest) hospitalizations per 100,000.

Considering state-specific hospitalization rates over time, not all states had the same peaks or magnitudes of peaks. Figure 3 shows the histogram across the study period for the ten states with the highest cumulative hospitalization rates from May 2020 through April 2021. From these example states, we were able to observe differences in the time trends between states regarding the timing and number of peaks. States including Texas, Nevada, Alabama, Arizona, and Tennessee have two peaks; however, they differed by timing and magnitude of the peaks. In contrast, Nebraska, Kansas, Virginia, Missouri, and Pennsylvania experienced only one major peak, which also differed by timing and magnitude. Hospitalization rates per 100,000 population from the final output model overtime are provided below in Figure 3.

**Figure 3:**
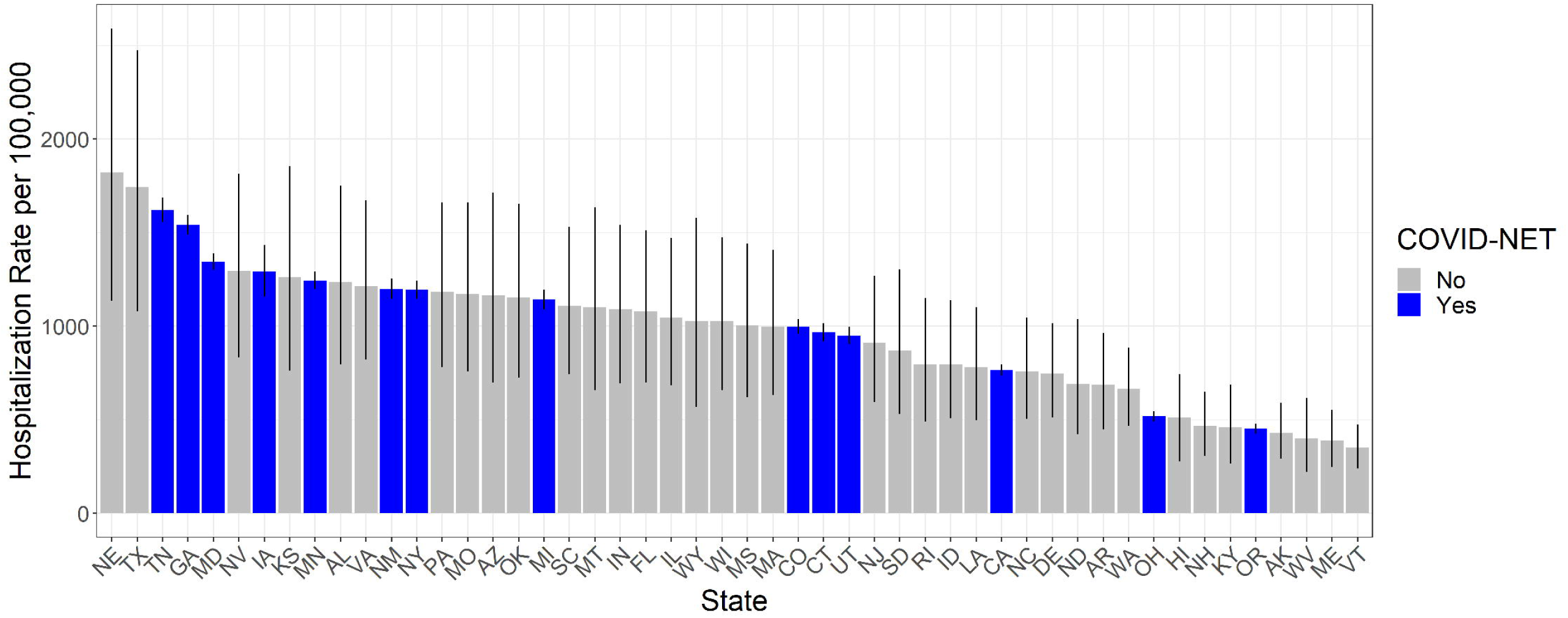
Hospitalization rates per 100,000 population over time for the 10 states with the highest cumulative hospitalization rates from May 2020 through April 2021.

To assess the sensitivity of the selected covariates, we ran the model using multiple combinations of the covariates, including those selected by LASSO method alone and those by spike and slab method alone. Hospitalization estimates did not vary greatly overall or by age depending on covariate combinations, and were almost 100% consistent between LASSO alone, Spike and Slab alone, and when both were used, which are the covariates used in the final model for each age group. To validate the final model, we compared the observed COVID-NET hospitalization rates to the final model’s estimated hospitalization rates. The rates are higher from the final model. However, the trends overtime and by age group follow the observed, input rates (S Figure 1). The supplementary images are a plot of each state comparing observed (input), estimated (final model), and extrapolated monthly hospitalization rate in the leave-one-state out analysis, showing rates over time and by age group. Model median results for other states were mostly consistent whether the specific COVID-NET state was dropped or not. Almost all of the COVID-NET states’ extrapolated estimates, i.e. when dropped, had a 90% Credible Interval that included the observed (input) estimate and estimated (final model) rate. The older age groups were more consistent and had more overlap between estimates than the younger age groups in the leave-one-state out analysis. Finally, we compared our output with other hospitalization estimates and data. First, we compared our results to the Unified Hospital Timeseries data and data published on The COVID Tracking Project.^23,24^ Figure 4 shows comparison of hospitalization rate from each source over time. Our model has higher estimates than the published Unified Hospital Timeseries data but shows the same trends and includes the rates in our 90% Credible Intervals for a few months. Finally, we compared our results to the current published numbers from CDC on hospitalizations based on CDC’s case-based multiplier model.^25^ Our model’s output was lower than the estimates from CDC’s case-based multiplier model (S Table 2). From June 2020 to March 2021, our model estimated a cumulative incidence of 900.7 per 100,000 population whereas CDC’s case-based multiplier estimated 1,345.3 per 100,000 population.

**Figure 4:**
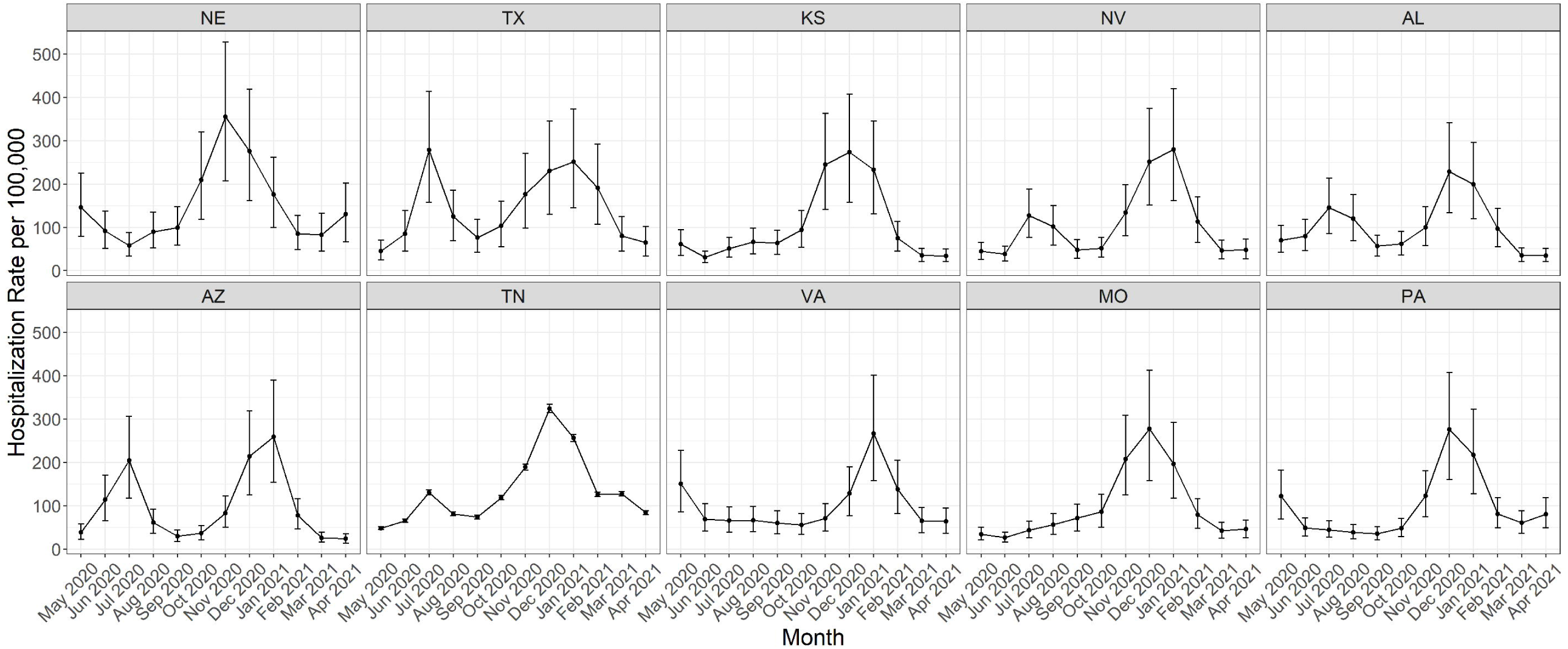
Comparison of hospitalization rates per 100,000 population over time from May 2020 through April 2021 from our final model output with 90% credible intervals, the Unified Hospital Timeseries data, and COVID Tracking.

## DISCUSSION

Overall, our method estimated 3,569,500 hospitalizations occurred in the United States from May 2020 through April 2021, with estimated rates varying by age group, state, and month. These estimates demonstrate the large burden of COVID-19 hospitalizations in the United States and provide visibility on variations in disease burden by age group, state, and time. As expected, the most severe burden of COVID-19 hospitalizations occurred among older age groups, specifically among people aged ≥65 years old. The largest peak in hospitalizations occurred in December 2020 and January 2021, aligning with the largest peak in reported case rates.^26^

Our approach to estimating the burden of COVID-19 hospitalization using long-term surveillance data has several benefits. First, we designed our model to build on an existing system that was initially started to track hospitalization for influenza and has expanded to capture other respiratory viruses including COVID-19. COVID-NET was built on a long-standing surveillance infrastructure that has been conducting surveillance for respiratory infections, including influenza and respiratory syncytial virus, for many years and is expected to continue monitoring COVID-19 hospitalization rates into the future.^27^ Our model calculated estimates of state-level hospitalization rates by month and age group, rather than assuming the 14 COVID-NET sentinel sites are representative of the United States. Each U.S. state has experienced the pandemic differently, and our models allow us to capture the variations in the number and magnitude of peaks, and state-specific trends in hospitalization rates. Further, using covariates to extrapolate data from the COVID-NET sites to the rest of the United States provides useful information to understand state-level differences in hospitalization. The covariates add information to the input hospitalization rates to then create a better story to the states it extrapolates to. This model helps preserve notable differences in the epidemiology of COVID-19 between states.

For initial estimates of COVID-19 hospitalizations, CDC used a multiplier method using nationally-notifiable case report data and assumptions for under-detection of confirmed cases.^5^ When we compared our model’s output to CDC’s case-based multiplier model during time periods that overlapped, we found that our model generated more conservative estimates of hospitalization. When comparing estimates by age group, months, and HHS regions, specific differences are highlighted. Our model had much lower estimates of hospitalization rates per 100,000 for the 0-17 year old age group, 210.7 for CDC’s multiplier model and 64.8 for ours, and 65 years old and above age group, 4,401.7 for CDC’s multiplier model and 2,794.3 for ours, while the other age groups were only slightly lower (Supplementary table 2). Also, our February through March estimate and HHS regions 2 and 9 were much lower. However, our model had higher estimates for a few HHS regions compared to CDC’s case-based multiplier estimates. Our method has several advantages over CDC’s case-based multiplier method. First, the case report data used was often incomplete for hospitalization status and relied on imputation of hospitalization status. In our method, the input hospitalization data were from a surveillance system that actively identified laboratory-confirmed COVID-19 hospitalizations. This may account for the differences observed in the hospitalization estimates between the models. Imputation could lead to more hospitalizations than those counted from the surveillance system. A second difference was that CDC’s case-based multiplier method adjusted reported cases for factors that influenced case detection, including health care seeking behaviors and testing practices, which were not available at the state level. Therefore, they adjusted and estimated at the HHS region level rather than the state level.

CDC’s case-based multiplier model relies on COVID-19 being a nationally notifiable disease and continued case reporting by states and jurisdictions which may not continue long-term. In contrast, our method relies on routine sentinel surveillance data and allows for extrapolation to places without data. Both the case report data and seroprevalence data used by Angulo et al.^6^ as the basis for their national COVID-19 disease burden estimates were data sources created to inform the pandemic response, but it is unclear how long these data will continue to be collected.

Although we utilized this method for estimating state-level hospitalization rates for COVID-19 in the United States from May 2020 through April 2021, our method can be adapted for different outcomes or measures of interest both domestically and in international settings. The main components needed are reliable surveillance data in enough areas to have diversity of disease occurrence and covariates that help explain the variation between all areas of extrapolation. There are surveillance systems set up that do not have complete coverage. For example, this approach was adapted from an analysis using a Bayesian Hierarchical model to extrapolate influenza yearly rates by country.^22^ This method provides an opportunity to leverage surveillance data and inform more accurate estimates of disease burden. Efforts to further expand the method to other levels of disease severity including infection, illness, or death are ongoing.

Our method also has some limitations. First, since our goal was to use routine surveillance data, our time frame for estimates began in May 2020 in states where we believe the surveillance systems were established and providing stable data after being set up in the early months of the pandemic. Therefore, we cannot estimate cumulative hospitalizations since the start of the pandemic. Second, we assume that COVID-NET captures all patients that were tested for COVID-19 and had a positive result. Although we adjust for testing practices, i.e., those not tested, we could be underestimating hospitalizations if the above assumption is not true and confirmed positives are not being reported. Third, we assumed that testing practices did not differ by states, except in Connecticut where testing practice data for COVID-NET sites was available. This assumption could result in either an over- or under-estimation of hospitalizations. Also, we assumed testing sensitivity for COVID-19 in COVID-NET was 0.885, which can lead to an over or under estimation of hospitalizations depending on true sensitivity. Fourth, our method assumes that the COVID-NET sites are representative of the entire state. In some states, such as Maryland, COVID-NET includes all counties; in other states, such as Iowa, it includes only one county. Although the model accounted for uncertainty and variability between states, we are still limited by representativeness within a state between the COVID-NET site and the truth of the entire state. As a result, our model may be under- or over-estimating hospitalizations at the state level for COVID-NET states depending on how well the particular catchment area reflects COVID-19 activity in the state. Fifth, our method assumes that COVID-NET states capture enough diversity across the nation to extrapolate data to all states, which may not be true. Although the 14 states from COVID-NET vary in many ways, we cannot be sure that they cover the variation in COVID-19 hospitalizations, including variations of thing that may impact hospitalizations like mitigation strategies and vaccination rates. For example, we could not extrapolate to Washington D.C. or New York City appropriately due to the extreme variation between a state and a purely metropolitan city. Sixth, although the covariates are meant to inform the extrapolation, the covariates are limited by the quality, completeness, and availability of the data. There could be vital information around COVID-19 hospitalization rates that are missing, such as other chronic conditions, underlying risk factors in the population, mitigation measures, and vaccination rates. Although our model has time-varying covariates that describe the COVID-19 impact in each state, including percent positive, percent COVID-19 deaths, and hospital capacity covariates, vaccination rates were not included so we may be under- or over-estimating age groups and states based on potential unaccounted variation from the correlation to vaccination rates. Another limitation is the wide credible intervals. Median estimates from the model’s output distributions of hospitalizations seems to be reasonable through our sensitivity, validation, and comparison analysis, but the 90% credible intervals are wide for some of the states where extrapolation was carried out. This limits precision of true hospitalizations and inference of medians presented. Finally, since we run a different model for each age group, we are limited in interpretation of hospitalization estimates by month and state since combining models’ output may underestimate variability and does not capture correlations between age groups. Although we calculated hospitalizations by month and state, combined variance is unknown so credible intervals may be wider than reported.

In conclusion, we estimated that about 4 million hospitalizations due to COVID-19 occurred in the United States from May 2020 through April 2021. As COVID-19 continues to circulate and cause illness, it will be important to develop a sustainable method to continue to estimate disease burden of COVID-19 that can account for regional variation in timing and incidence of disease activity, changes in detection and reporting of COVID-19, and that utilizes ongoing surveillance data. With an unknown future of COVID-19, burden estimates will continue to be needed. Having a burden estimation method that uses a sentinel surveillance system ensures we will have the ability to create burden estimates despite changes in case data reporting. Knowing disease burden helps us understand vaccine averted burden, post COVID-19 conditions, and more important public health research. Our method leverages routine surveillance data that are expected to continue after the pandemic and a Bayesian Hierarchical modelling approach as a novel way to continue estimating COVID-19 hospitalization. The model offers an approach that will be useful not only to COVID-19 hospitalization estimations but to other levels of the disease burden pyramid, including infections and deaths.

## Data Availability

Data will not be made available online.

## Authors’ Contributions

AC, ADI, HHC, NNP, MG, MS, and CR were involved in the study conceptualization. AC, ADI, NNP, MG, MW, and CR analyzed the primary and input data. AC and HHC coded the model and AC was responsible for running all model analyses. AC, ADI, NNP, MG, MS, FPH, MW, and CR validated the data used in the analysis. AC and CR led the initial drafting of the manuscript. All authors contributed to ideas for analysis and manuscript edits and subsequent drafts. All authors had access to the underlying data, have seen and approved of the final text, and were responsible for the decision to submit.

## Abbreviations

BRFSS: Behavioral Risk Factor Surveillance System
CDC: Centers for Disease Control and Prevention
CKD: chronic kidney disease
COPD: chronic obstructive pulmonary disease
COVID-19: Coronavirus Disease 2019
COVID-NET: Coronavirus Disease 2019-Associated Hospitalization Surveillance Network
CrI: Credible Interval
FluSurv-NET: Influenza Hospitalization Surveillance Network
HHS: Department of Health and Human Services
ICU: intensive care unit
LASSO: Least Absolute Shrinkage and Selection Operator
MCMC: Markov chain Monte Carlo
NCHS: National Center for Health Statistics
NNDSS: National Notifiable Disease Surveillance System
NVSS: National Vital Statistics System
SARS-CoV-2: Severe Acute Respiratory Syndrome Coronavirus 2

## Supplement

**Table S1:**
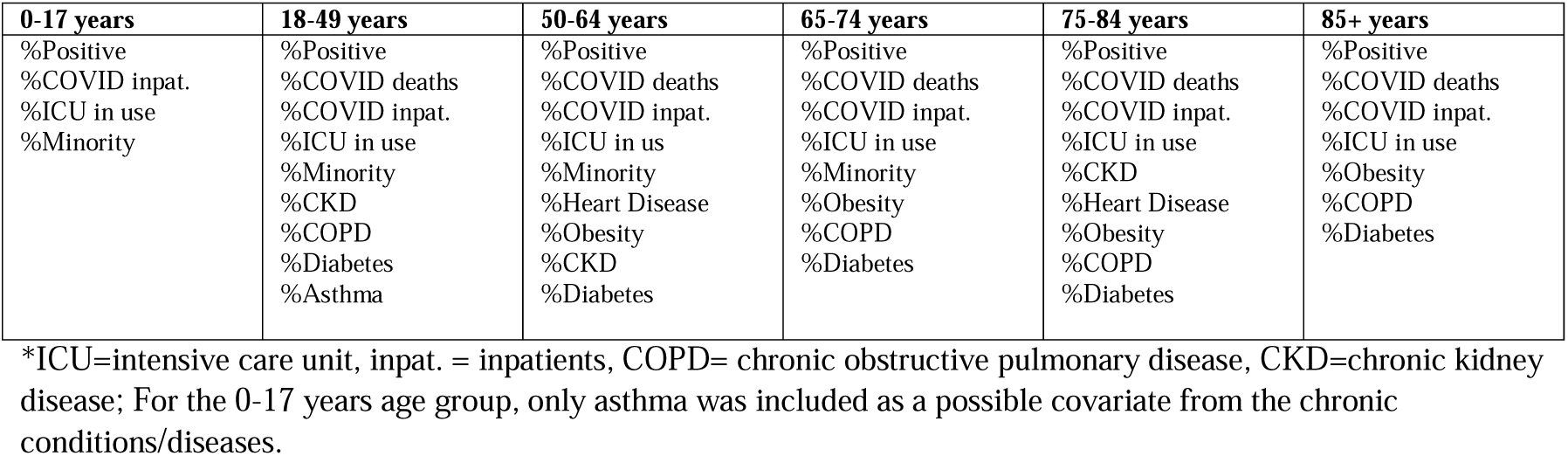
Covariates selected for each model by age group by selection methods out of all covariates.

| 0-17 years | 18-49 years | 50-64 years | 65-74 years | 75-84 years | 85+ years |
| --- | --- | --- | --- | --- | --- |
| %Positive<br>%COVID inpat.<br>%ICU in use<br>%Minority | %Positive<br>%COVID deaths<br>%COVID inpat.<br>%ICU in use<br>%Minority<br>%CKD<br>%COPD<br>%Diabetes<br>%Asthma | %Positive<br>%COVID deaths<br>%COVID inpat.<br>%ICU in use<br>%Minority<br>%Heart Disease<br>%Obesity<br>%CKD<br>%Diabetes | %Positive<br>%COVID deaths<br>%COVID inpat.<br>%ICU in use<br>%Minority<br>%Obesity<br>%COPD<br>%Diabetes | %Positive<br>%COVID deaths<br>%COVID inpat.<br>%ICU in use<br>%CKD<br>%Heart Disease<br>%Obesity<br>%COPD<br>%Diabetes | %Positive<br>%COVID deaths<br>%COVID inpat.<br>%ICU in use<br>%Obesity<br>%COPD<br>%Diabetes |

\*ICU=intensive care unit, inpat. = inpatients, COPD= chronic obstructive pulmonary disease, CKD=chronic kidney disease; For the 0-17 years age group, only asthma was included as a possible covariate from the chronic conditions/diseases.

**Table S2:** Comparison of hospitalization estimates between Bayesian model and case-based multiplier model by age group, months, and HHS regions, including distribution of hospitalization for each group from June 2020 through March 2021.

| Group | Final Bayesian Model |  | Case-Based Multiplier Model |  |
| --- | --- | --- | --- | --- |
|  | Hospitalization Count<br>(% of Group) | Hospitalization Rate per<br>100,000 | Hospitalization Count<br>(% of Group) | Hospitalization Rate per<br>100,000 |
| **Age Groups** |  |  |  |  |
| Ages 0-17 | 51,400 (1.6) | 64.8 | 167,085 (3.6) | 210.7 |
| Ages 18-49 | 775,700 (24.9) | 516.4 | 1,124,575 (24.1) | 748.6 |
| Ages 50-64 | 784,200 (25.1) | 1248.3 | 1,185,968 (25.5) | 1887.9 |
| Ages 65+ | 1,508,100 (48.3) | 2794.3 | 2,181,343 (46.8) | 4041.7 |
| **Months** |  |  |  |  |
| June - July | 458,800 (14.7) | 138.2 | 572,148 (12.3) | 172.4 |
| August - September | 359,400 (11.5) | 109.7 | 404,311 (8.7) | 123.4 |
| October - November | 603,600 (19.3) | 184.3 | 1,008,710 (21.7) | 308.0 |
| December - January | 1,207,300 (38.7) | 368.6 | 1,585,383 (34.0) | 484.0 |
| February - March | 490,400 (15.7) | 146.4 | 1,088,419 (23.4) | 325.0 |
| **HHS Regions** |  |  |  |  |
| Reg1 | 169,200 (5.4) | 1139.8 | 148,947 (3.2) | 1003.3 |
| Reg2 | 261,700 (8.4) | 923.5 | 797,480 (17.1) | 2814.4 |
| Reg3 | 259,600 (8.3) | 691.1 | 327,607 (7.0) | 872.2 |
| Reg4 | 683,000 (21.9) | 1020.8 | 625,422 (13.4) | 934.7 |
| Reg5 | 421,600 (13.5) | 802.4 | 815,382 (17.5) | 1551.9 |
| Reg6 | 572,600 (18.4) | 1340.4 | 672,674 (14.4) | 1574.7 |
| Reg7 | 168,400 (5.4) | 1190.7 | 148,695 (3.2) | 1051.6 |
| Reg8 | 102,900 (3.3) | 616.3 | 104,509 (2.2) | 626.1 |
| Reg9 | 405,700 (13) | 791.1 | 940,347 (20.2) | 1833.5 |
| Reg10 | 74,800 (2.4) | 521.3 | 77,908 (1.7) | 542.9 |
| **Total** | 3,119,400 | 900.7 | 4,658,971 | 1345..3 |

**Figure S1:**
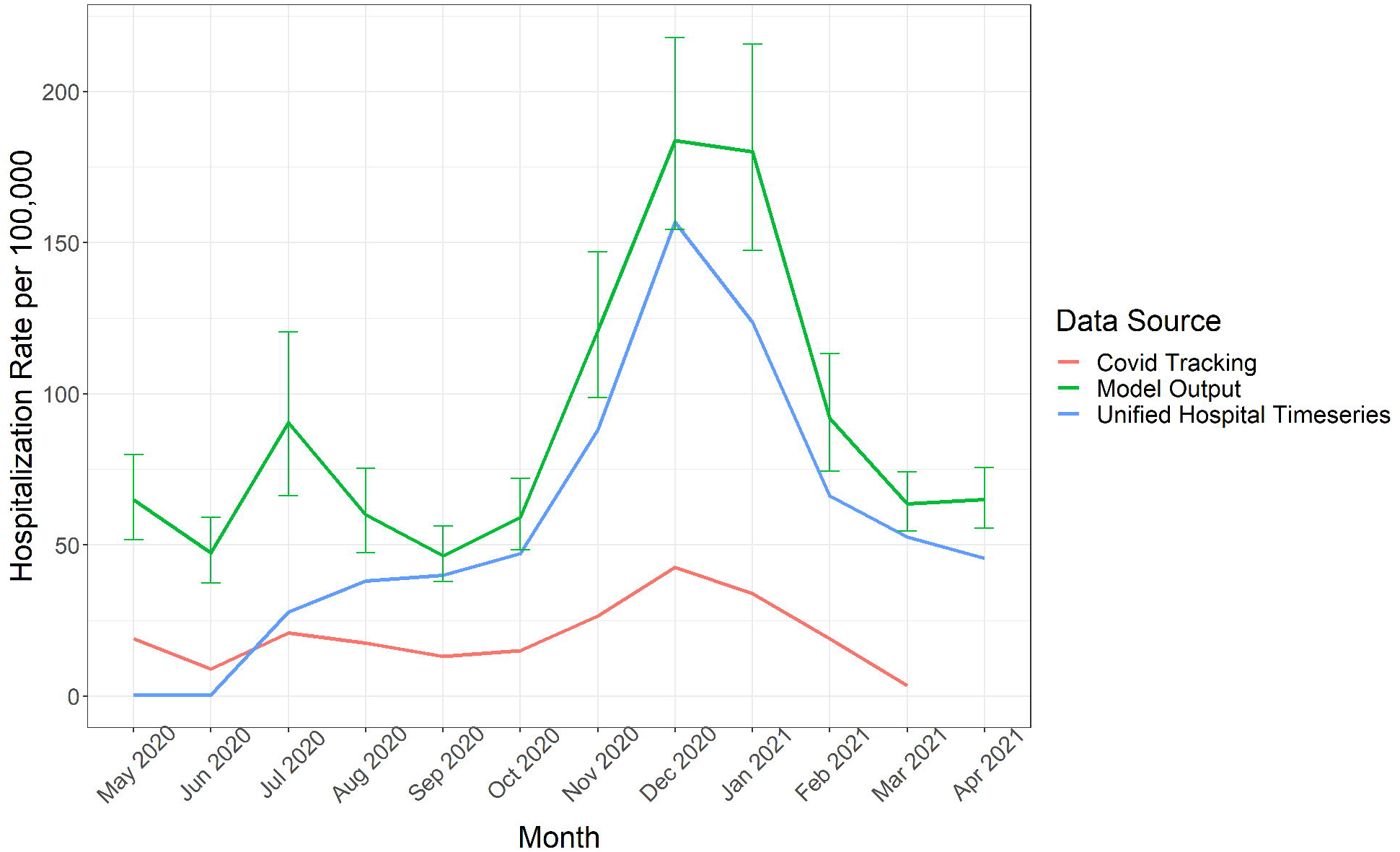
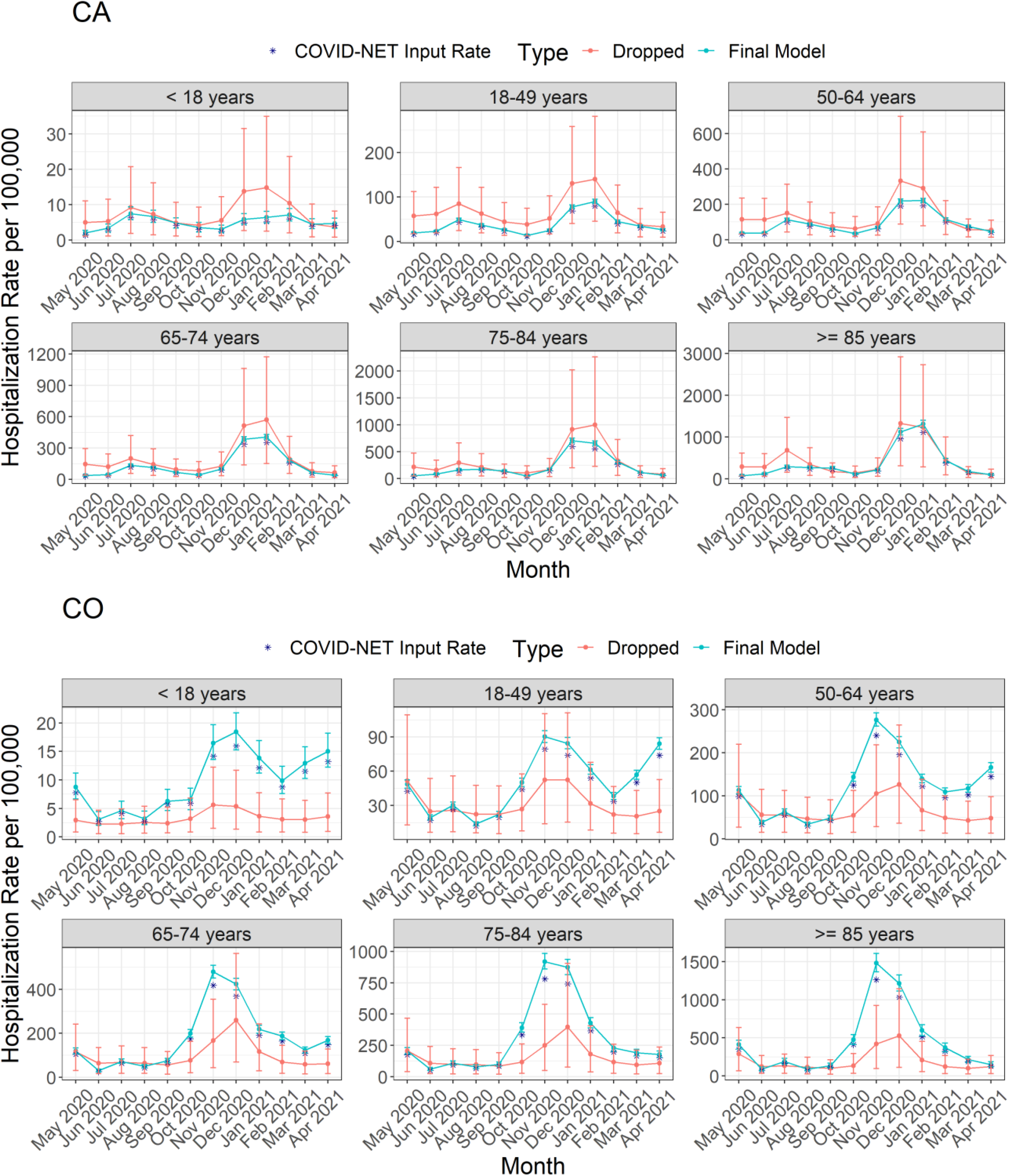
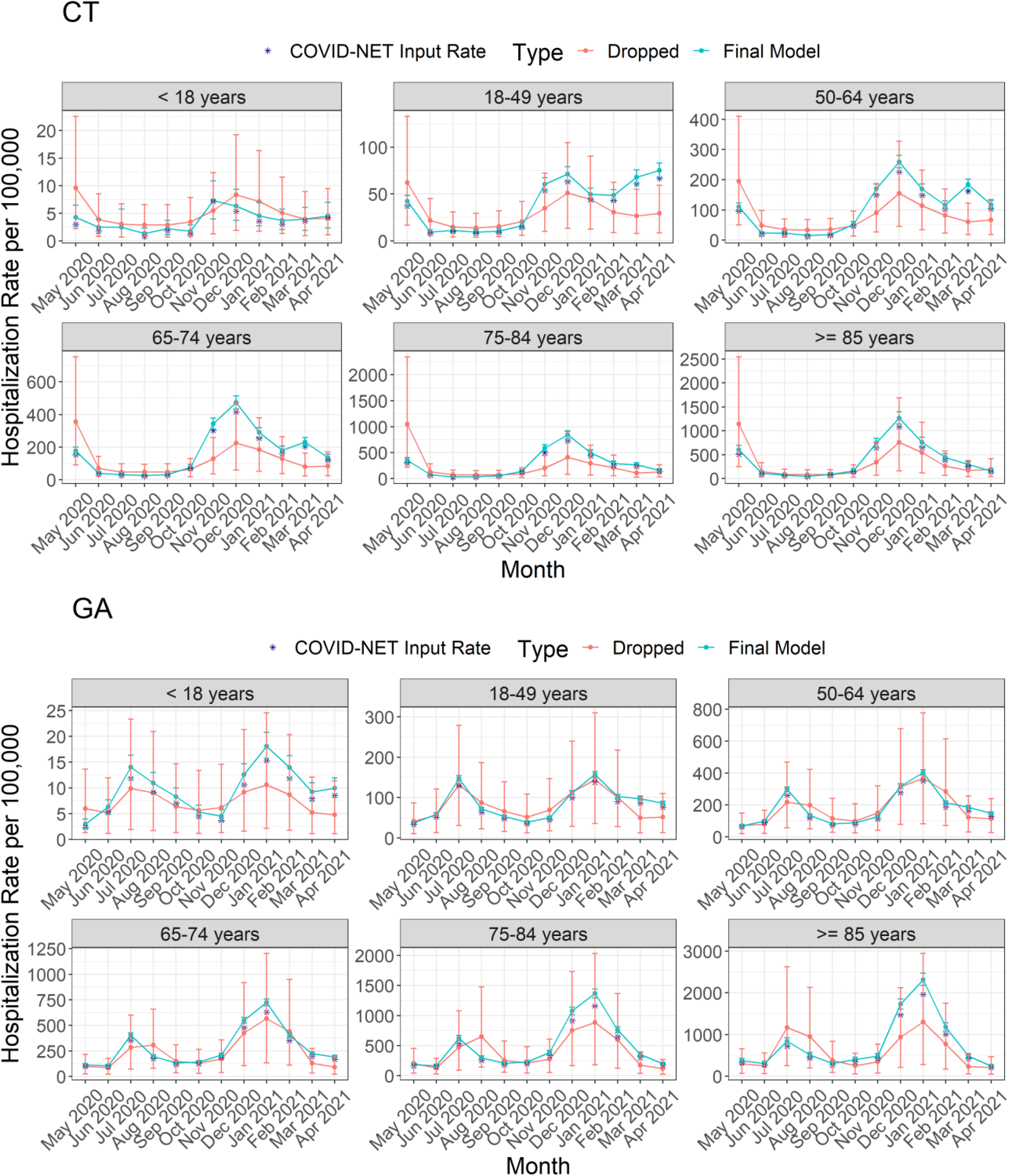
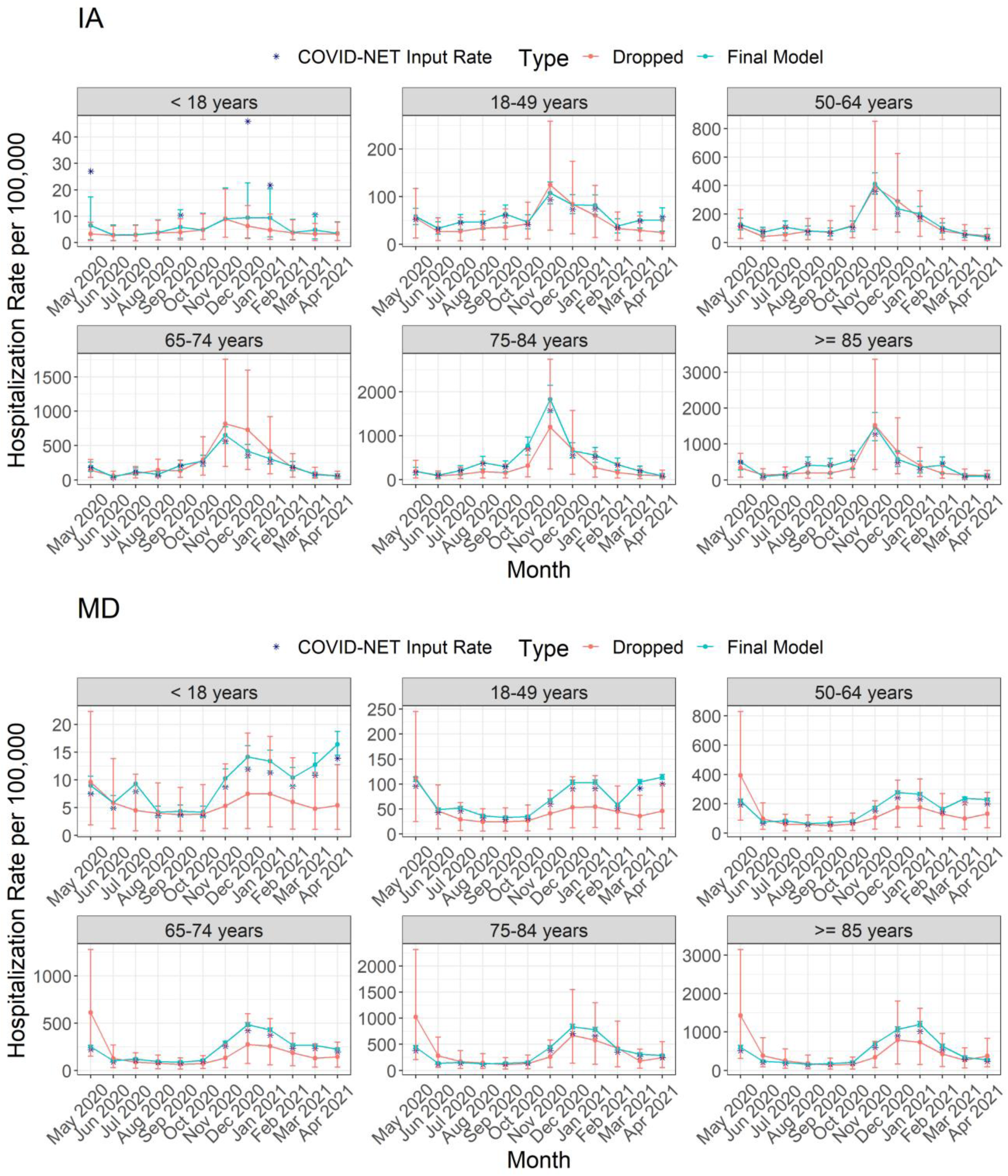
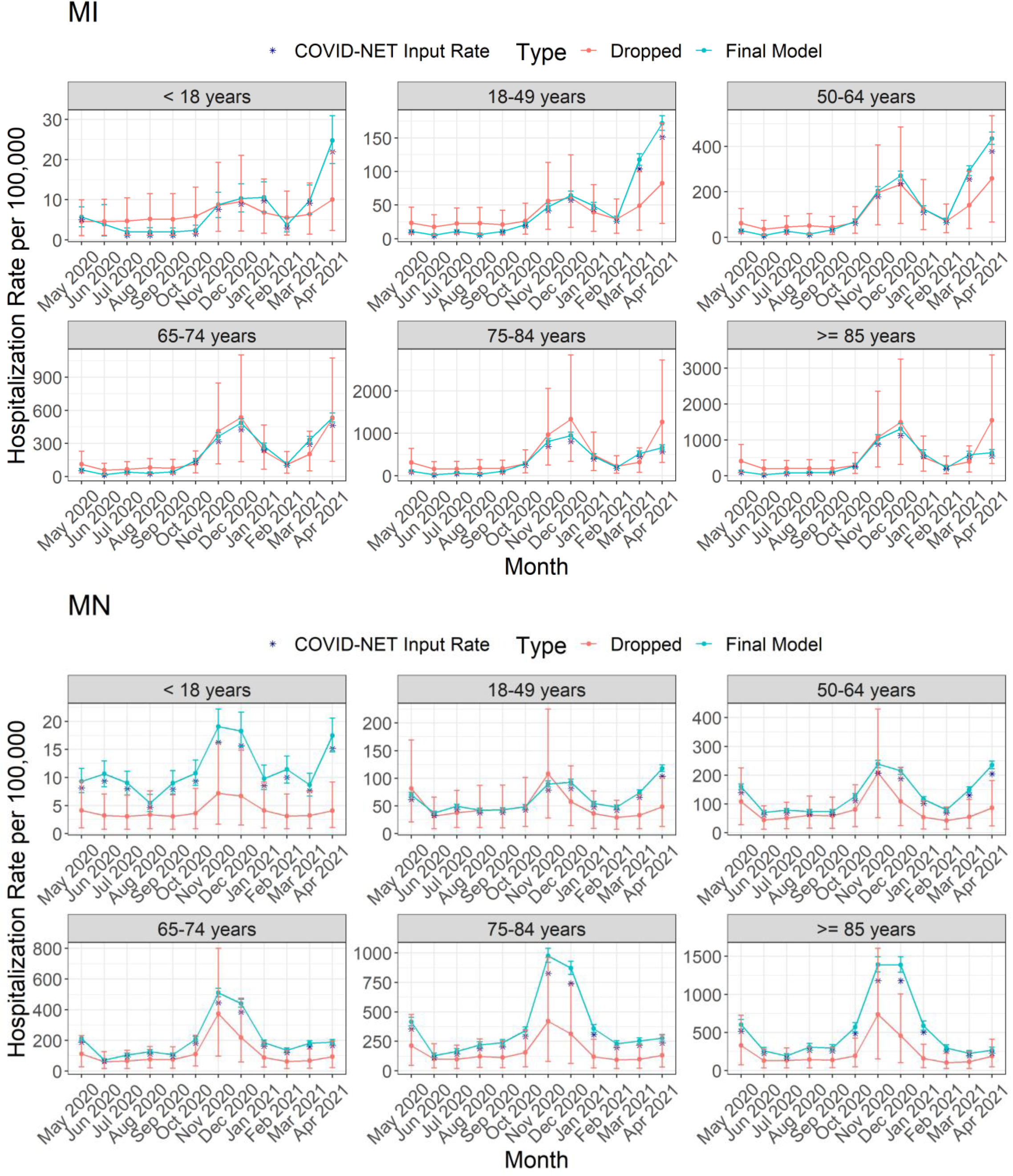
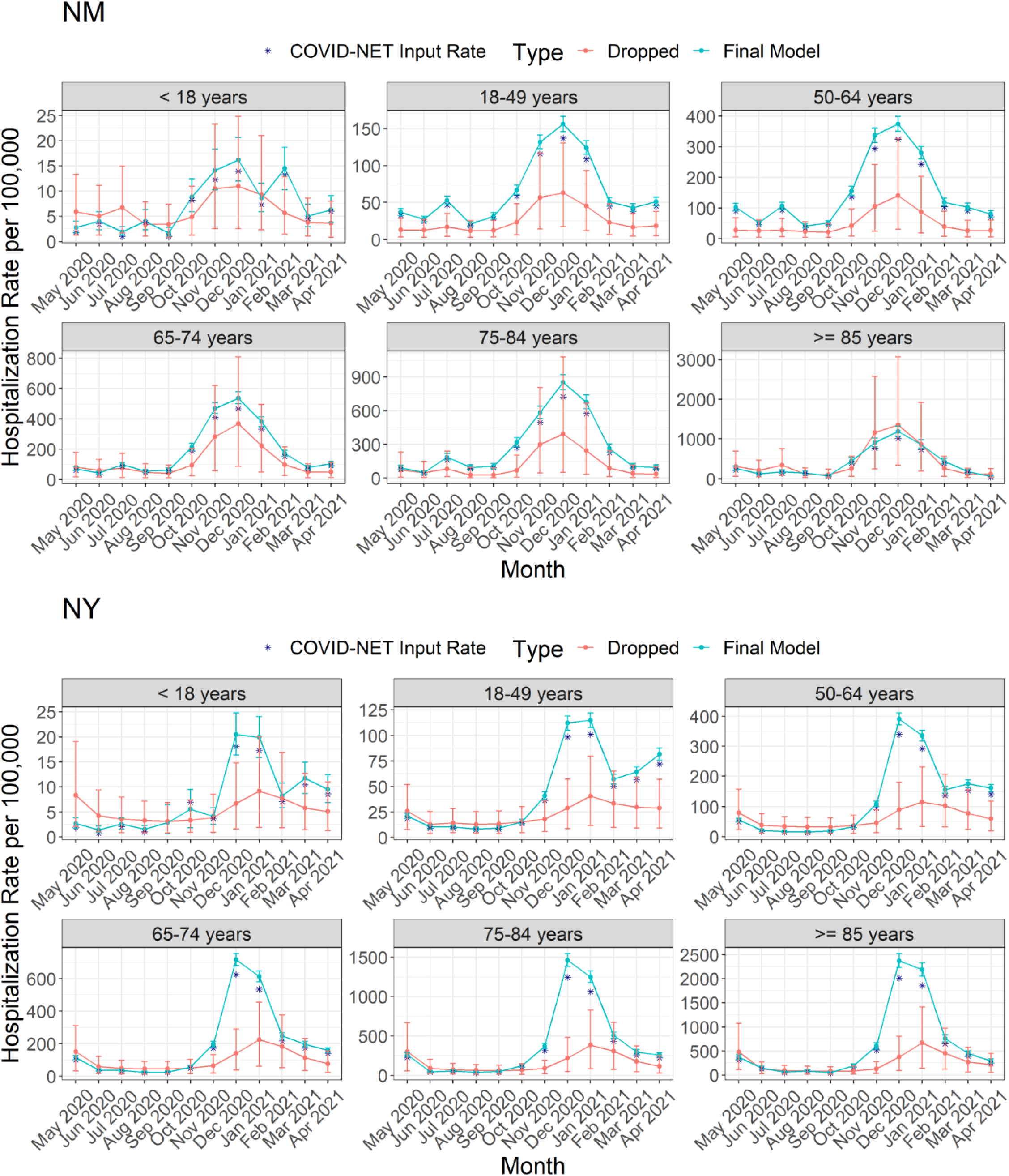
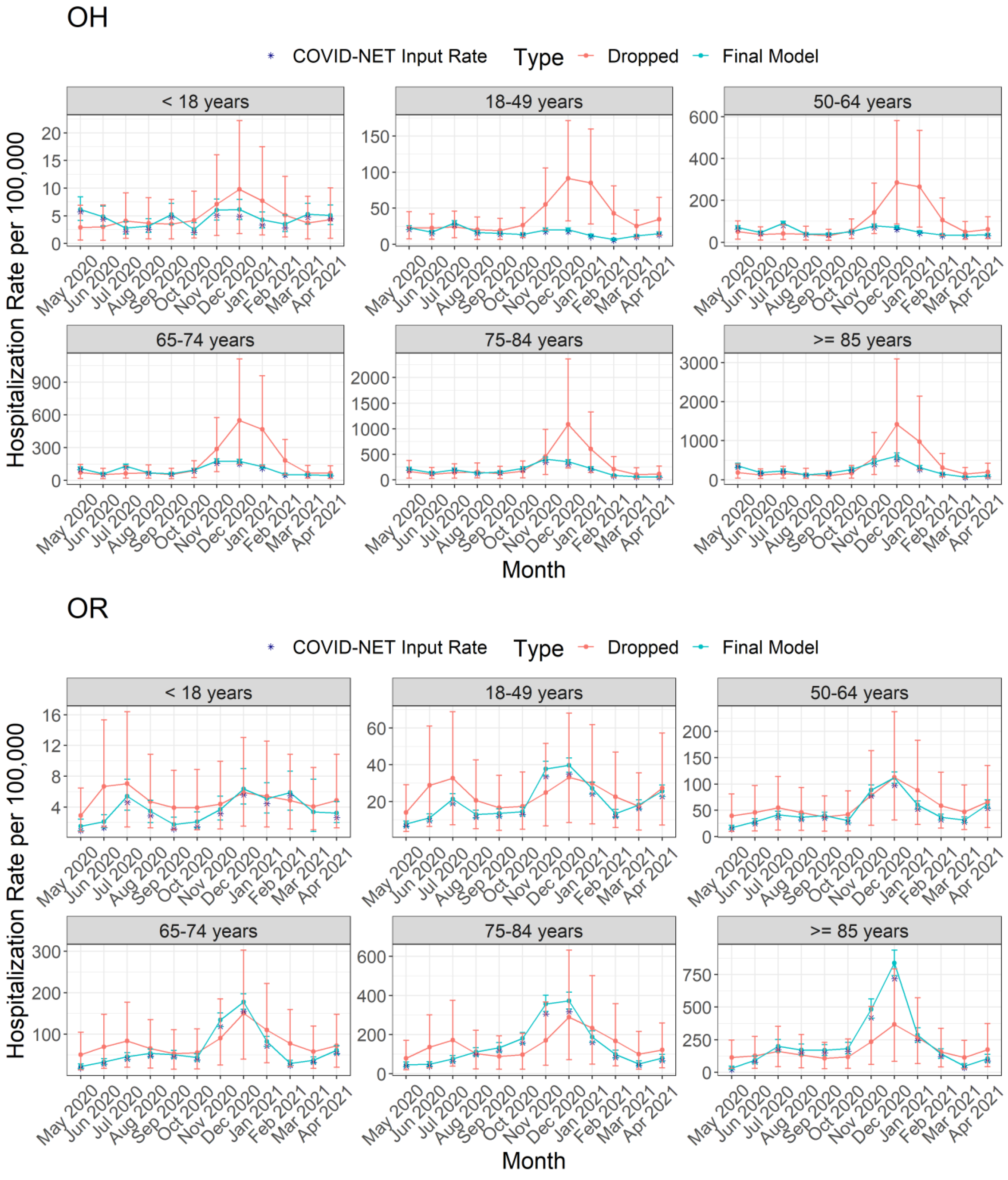
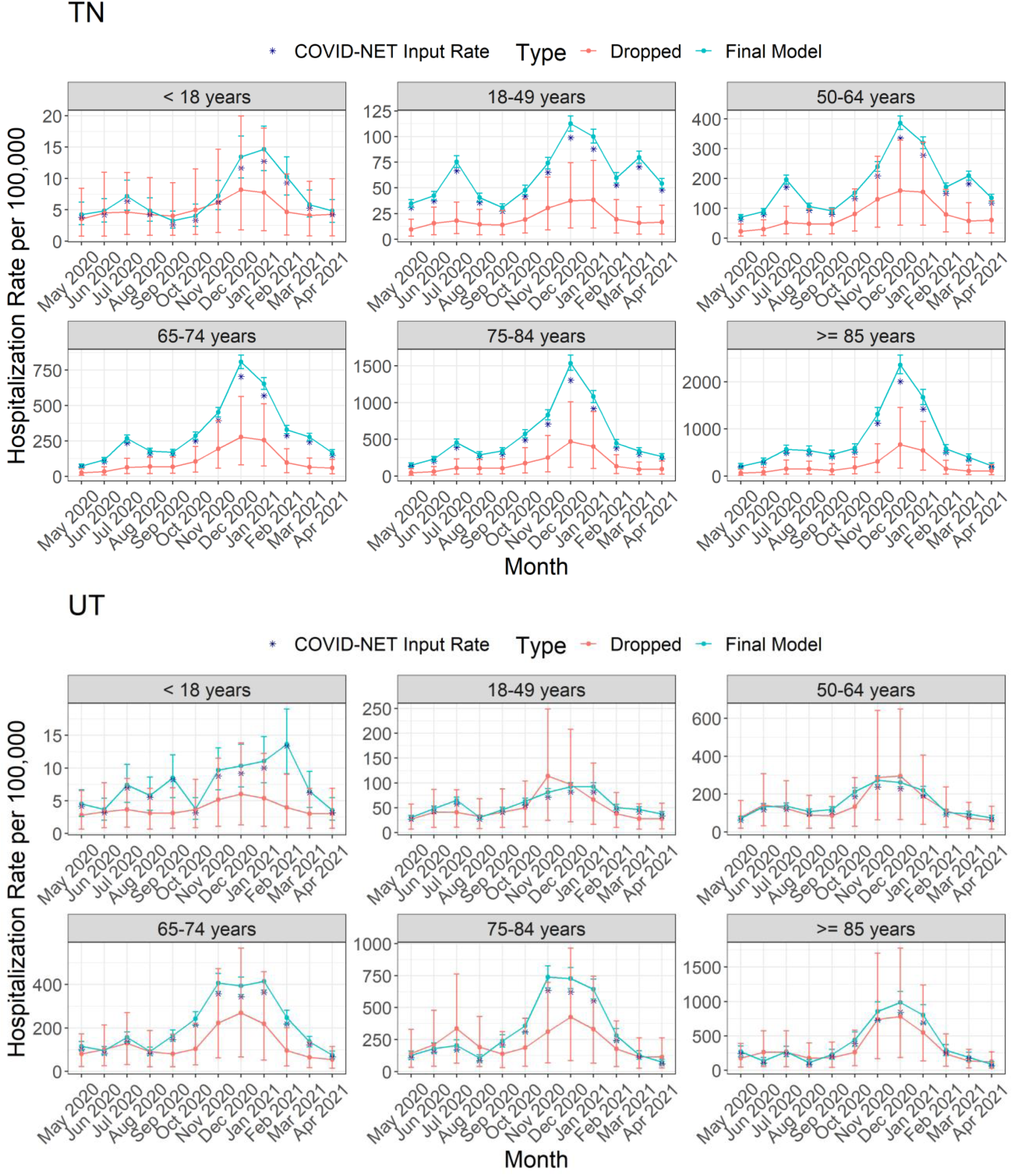
Comparison of hospitalization rates per 100,000 population and 90% credible intervals (error bars) from model by age group for each state in COVID-NET showing observed rate (COVID-NET Input Rate), estimated rate (Final Model), and extrapolated rate (Dropped). Y-axis limits adjust to the unique minimum and maximum rate for each age group.

